# Data driven phenotyping and COVID-19 case definitions: a pattern recognition approach

**DOI:** 10.1101/2021.04.30.21256219

**Authors:** George D. Vavougios, Christoforos Konstantatos, Pavlos-Christoforos Sinigalias, Sotirios G. Zarogiannis, Konstantinos Kolomvatsos, George Stamoulis, Konstantinos I. Gourgoulianis

**Affiliations:** Department of Computer Science and Telecommunications, University of Thessaly, Papasiopoulou 2–4, Galaneika, Lamia 35131, Greece; Department of Respiratory Medicine, Faculty of Medicine, School of Health Sciences, University of Thessaly, Biopolis, Larissa 41500, Greece; Department of Business Administration, University of Patras, University Campus – Rio, Patras 26504, Greece; Department of Mechanical Engineering and Aeronautics, University of Patras, Greece; Department of Physiology, Faculty of Medicine, School of Health Sciences, University of Thessaly, BIOPOLIS, Larissa 41500, Greece; Department of Electrical and Computer Engineering, University of Thessaly, 37 Glavani – 28^th^ October Str, Deligiorgi Building, 4th floor, Volos 38221, Greece

**Keywords:** COVID-19, Pattern recognition, phenotypes, epidemiology, comorbidity, big data

## Abstract

**Introduction:** COVID-19 has pathological pulmonary as well as several extrapulmonary manifestations and thus many different symptoms may arise in patients. The aim of our study was to determine COVID-19 syndromic phenotypes in a data driven manner using survey results extracted from Carnegie Mellon University’s Delphi Group.

**Methods:** Monthly survey results (>1 million responders per month; 320.326 responders with positive COVID-19 test and disease duration <30 days were included in this study) were used sequentially in identifying and validating COVID-19 syndromic phenotypes. Logistic Regression Weighted Multiple Correspondence Analysis (LRW-MCA) was used as a preprocessing procedure, in order to weight and transform symptoms recorded by the survey to eigenspace coordinates (i.e. object scores per case / dimension), with a goal of capturing a total variance of > 75%. These scores along with symptom duration were subsequently used by the Two Step Clustering algorithm to produce symptom clusters. Post-hoc logistic regression models adjusting for age, gender and comorbidities and confirmatory linear principal components analyses were used to further explore the data. The model created from 66.165 included responders in August, was subsequently validated in data from March – December 2020.

**Results:** Five validated COVID-19 syndromes were identified in August: 1. Afebrile (0%), Non-Coughing (0%), Oligosymptomatic (ANCOS) 2. Febrile (100%) Multisymptomatic (FMS) 3. Afebrile (0%) Coughing (100%) Oligosymptomatic (ACOS), 4. Oligosymptomatic with additional self-described symptoms (100%; OSDS) and 5. Olfaction / Gustatory Impairment Predominant (100%; OGIP).

**Discussion:** We present 5 distinct symptom phenotypes within the COVID-19 spectrum that remain stable within 9 – 12 days of first symptom onset. The typical febrile respiratory phenotype is presented as a minority among identified syndromes, a finding that may impact both epidemiological surveillance norms and transmission dynamics.

## 1. Introduction

Since its emergence, COVID-19 has conceptually evolved from a viral pneumonia to a multisystem disease with insidious onset and diverse outcomes (1). As additional cases caused a shift in case definitions, big data and detailed symptom indexing arose as a necessity toward guiding evidence-based medicine and preventing severe outcomes (2). An intrinsic perturbation in using expert-based definitions is inherent bias (i.e. as this is a hypothesis- or observation-driven approach), and an expected lack of recognition of fringe cases or spectrums, that may however be deemed as such due to their underrepresentation within a given cohort.

We were the first to phenotype comorbidity in obstructive sleep apnea (OSA) using a combination of preclustering principal component analysis to identify latent structures within our datasets, and subsequently map them using the TwoStep Cluster algorithm (3). The methodology we proposed has been adopted by other research groups (4 – 8) and successfully implemented in other disease models, enabling the ad hoc development of diverse phenotyping concepts (9 – 12).

Data-driven recognition of COVID-19 phenotypes will allow an unbiased mapping of the global clinical spectrum, along with identifying susceptibility and severity clusters that demand a significantly different clinical approach. Furthermore, these data-driven phenotypes would enable the design of clinical studies and enhance outcome design and evaluation, producing efficacious, bias-free treatments and interventions.

Therefore, the specific aims of the study were the following:

1. The use of reported symptoms to identify latent structures via categorical PCA and dimension reduction approaches, using data from the COVID-19 Delphi Facebook study.
2. To scrutinize the previously created latent structures as potential COVID-19 phenotypes or phenotyping parameters via TSC and artificial intelligence-based classification.

## 2. Materials and methods

### 2.1 Study Population

Data for this study were extracted from the COVID-19 Symptom study, based on symptom surveys run by the Delphi group at Carnegie Mellon University (CMU). Initially, Facebook selects a random sample among its users in the United States. The users are then presented with the option to participate in the study. In turn, participation entails the administration of the surveys iteration, and covers data on COVID-19-like symptoms, behavioral, mental health, and economic parameters, as well as estimates on the impact of the pandemic on the responders daily life. Individual, anonymized survey responses are stored in CMU’s servers and made accessible to healthcare professionals under a project-specific data use agreement. Approximately 50,000 responders participate in the study per day, with monthly survey results comprising more than 1 million responders per month.

A detailed overview of the COVID-19 symptom study, its conception and evolution are available from: https://cmu-delphi.github.io/delphi-epidata/symptom-survey/

#### 2.1.1. Inclusion and exclusion criteria

In this study, we included responders with a certain COVID-19 status, i.e. having answered either “Yes” or “No” in the corresponding item of the survey. An indicative item structure from Wave 4 of the study is the following:

- *B10 – Have you been tested for coronavirus (COVID-19) in the last 14 days?*
- *B10a – Did this test find that you had coronavirus (COVID-19)?*

A positive COVID-19 status was assigned to responders answering “Yes” in B10a, whereas a negative COVID-19 status was assigned to responders answering “No” in the same item. Responders that answered “I do not know” in item B10a were excluded from further analyses.

As a subsequent exclusion criterion, we employed a cutoff of ≤ 30 days in symptom duration. This cutoff was selected on the premises of an approximate “return to wellness”, estimated to occur at 14 – 21 days for 65% of patients a positive outpatient test result, in a recent report by a CDC (13). The purpose of this cut-off was to simultaneously include COVID-19 patients with longer duration illness and exclude symptom durations unlikely to be attribute to COVID-19 manifestations such as i.e. 60 days or more. In order for our approach to be forward and backward compatible, we opted to use the very first incarnation of symptoms attributed to COVID-19 and captured by the survey (i.e. the first wave). As such, a core of the 13 first symptoms recorded by the survey would remain the same, regardless of future additions.

## 3. Statistical Analysis

### 3.1. Logistic probability case scoring

The logistic regression model extracted from August’s data was subsequently used to score subsequent cases per month. Let us consider a model where *x*_1_,*x*_2_,…, *x*_*n*_ are symptoms (used as predictors) of the binary response variable *C (1=COVID-19 positive, 0=COVID-19 negative)*, which represents COVID-19 status. The log-odds λ of the probability *p* of *C* = 1 can be presented as follows:

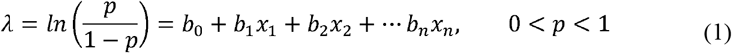

where *b*_1_,*b*_2_,…,*b*_*n*_ are the model parameters, *b*_0_ is the constant and 1– *p* is the probability of *C*= 0. The exponent of λ retrieves the odds for each case:

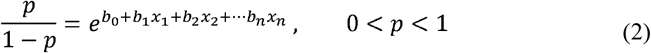

where *e* is Napier’s number. The probability *p* can then be expressed as follows:

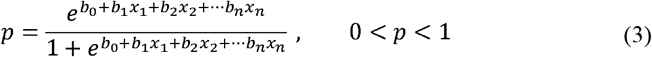

To generate our model, we calculate each *b*_*i*_, where *i* ⋲ [1… *n*] for each variable *x*_*l*_ where *i*⋲ [1 … *n*]. For each predictor (i.e., symptom) *x*_*l*_, the odds ratio calculated by logistic regression (i.e. the exponent of *b*_*l*_) is used as a corresponding weight in a subsequent multiple correspondence analysis. Finally, the probability *p* is used to score each case.

### 3.2. Pattern recognition via Multiple Correspondence Analysis

Symptom data were used by combining a dimension reduction technique and cluster analysis algorithm, as previously described (9,13). Object scores derived from a multiple correspondence analysis (MCA) of symptom data were subsequently used as input variables for the cluster analysis, along with symptom duration on COVID-19 positive responders (9,14). The optimal number of MCA-derived dimensions was determined based on achieving a total variance (i.e. cumulative variance per dimension) of > 70% (15). MCA and MCA-preprocessing prior to cluster analysis are techniques that allow the identification of latent patterns within a population, based on a set of nominal response variables (16); OR-weighting of the input variables is used here as a ranking scheme based on their association with COVID-19 positive responders vs. COVID-19 negative responders.

### 3.3. Two-Step Clustering and Phenotype Extraction

OR-weighted MCA produced case-wise object scores (i.e. composite quantifications of symptoms per case) along with symptom duration were subsequently used by the Two Step Clustering (TSC) algorithm to produce symptom phenotypes. As we and others have previously demonstrated, TSC is well suited for the identification of latent phenotypes in a given population (9,16). In this study, the Log-likelihood was used as a distance measure, and the Bayesian Information Criterion (BIC) was used as the clustering criterion for the automatic determination of cluster number.

### 3.4. Phenotype Validation: Cross-sectional and Longitudinal aspects

The model that was created on the pilot analysis of 66.165 responders within the August dataset, was subsequently validated in monthly data ranging from March – December 2020. Specifically, the procedure was as follows:

a. The weights extracted from August’s responders were applied to an MCA based on symptom data recorded for each subsequent and preceding month’s responders.
b. Object scores were calculated for each responder.
c. Object scores and symptom duration per month were used in TSC.

### 3.5. Crossectional validation: Phenotypes vs. controls

Crossectional validation of the produced phenotypes essentially answers the question of whether a COVID-19 syndrome is associated with a positive COVID-19 test. For this purpose, Receiver Operator Characteristic (ROC) curves were used to determine the diagnostic accuracy of a symptom-based probability for each phenotype, when compared to controls. Specifically, ROC curves were fitted by the probability *p*_*i*_ extracted from the application of the logistic regression model derived from August’s data. Hence, *p*_*i*_ would be expressed as follows:

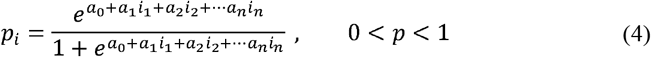

where *i*_1_,*i*_2_,…,*i*_*n*_ is the symptom per *i* – *th* month, and *a*_0_, *a*_1_, …, *a*_*n*_ are the *f* extracted from August’s dataset. Finally, COVID-19 status was used as a binary dependent variable for the ROC curve, and the area under curve (AUC) was calculated per month, for each cluster.

### 3.6. Longitudinal validation: Phenotype re-emergence and symptom invariance

The primary criterion for validation was the emergence of consistent phenotypes in at least one month other than August based on complete or quasi-complete symptom separation per phenotype. Essentially, this would translate in the identification of each phenotype based on the most salient or preclusive symptoms.

The secondary criterion was based on the hypothesis that re-emergent phenotypes would be furthermore identified based on non-salient, non-preclusive symptoms. To meet this criterion, frequency tables for each symptom were constructed per each month and phenotype.

For each symptom *S* reported on each month *M*, for a number of months *N*, we consider the mean, *µ*_*s*_:

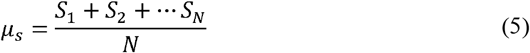

In order to assess symptom perseverance and their non-random contribution as patterns within each phenotype, we perform a normality test under the null hypothesis that the distribution of a symptom / month [*S*_1_,*S*_2_, … *S*_*N*_] is normal and therefore 95% of the observations lie within two standard deviations of *µ*_*s*_. The following concept has been previously used in face recognition algorithms (22). Here, we used the Shapiro – Wilk test of normality, and a p-value < 0.05 was considered statistically significant. Symptoms achieving below threshold p-values were considered variable for each corresponding phenotype. Correspondingly, longitudinal symptom invariance (SI) was described as follows:

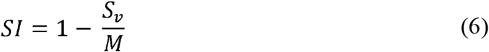

where *S*_*v*_ is the number of variant symptoms (defined by a Shapiro-Wilk p-value < 0.05) and M is the total number of symptoms. SI is equal to 1 (100%) when *S*_*v*_ =0 and SI is equal to 0 (0%) when *S*_*v*_ = *M*.

### 3.7. Post-hoc analyses

Associations between each phenotype and symptoms were determined via a combination of the *χ*^2^ test with adjusted standardized residuals (Standardized Pearson residuals). A *χ*^2^ test p-value < 0.05 and adjusted standardized residuals either greater than 1.96 or less than −1.96 was considered statistically significant. Associations among phenotypes and responder demographic and medical history characteristics (age group, gender, and comorbidity) were investigated via a logistic regression model.

### 3.8. Determination of Data-Driven Diagnostic Rules via Decision Tree analyses

In order to create a diagnostic rule per phenotype, we performed Decision Tree Analyses (DTA) via the Quick, Unbiased, Efficient, Statistical Tree (QUEST) algorithm (18). Decision tree analysis is a data mining technique that is implemented in order to create a classification scheme from a set of observations; in biomedical research, its main applications include the creation of data-driven diagnostic or predictive rules (19), (20).

QUEST was originally proposed by Loh and Shih (21). Let us consider cluster (i.e. phenotype) membership as a dependent nominal variable Y with J classes (equal to the number of phenotypes), and each symptom as a categorical (nominal) predictor *X*.

The decision tree expands by testing for the best predictors among the input, For each categorical predictor *X*, QUEST performs a chi-square test of independence at each node *n*, subsequently calculating the corresponding p-value:

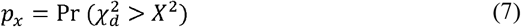

where *d* represents the degrees of freedom for 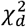, for a predictor *X* with *K*_*n*_ categories:

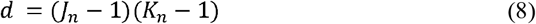

The growth procedure depends on establishing the best splitting predictor at each node based on the smallest p-value. Conversely, the “stopping” process is determined by several stopping criteria. In our study, the applied criterion was node purity, i.e. the complete separation of a predictor variable on a dependent variable class.

Each p-value < 0.05 was considered statistically significant. All analyses were performed SPSS version 24.0 (IBM, Chicago, Illinois, US).

## 4. Results

### 4.1. Study population

The total study population included 320.326 responders with a certain COVID-19 test status and disease duration <30 days (**Figure 1**). **Table 1** presents the study population’s demographics per month. **Supplementary Table 2** presents multinomial regression results per month and phenotype (**A1 – A5**).

**Figure 1.**
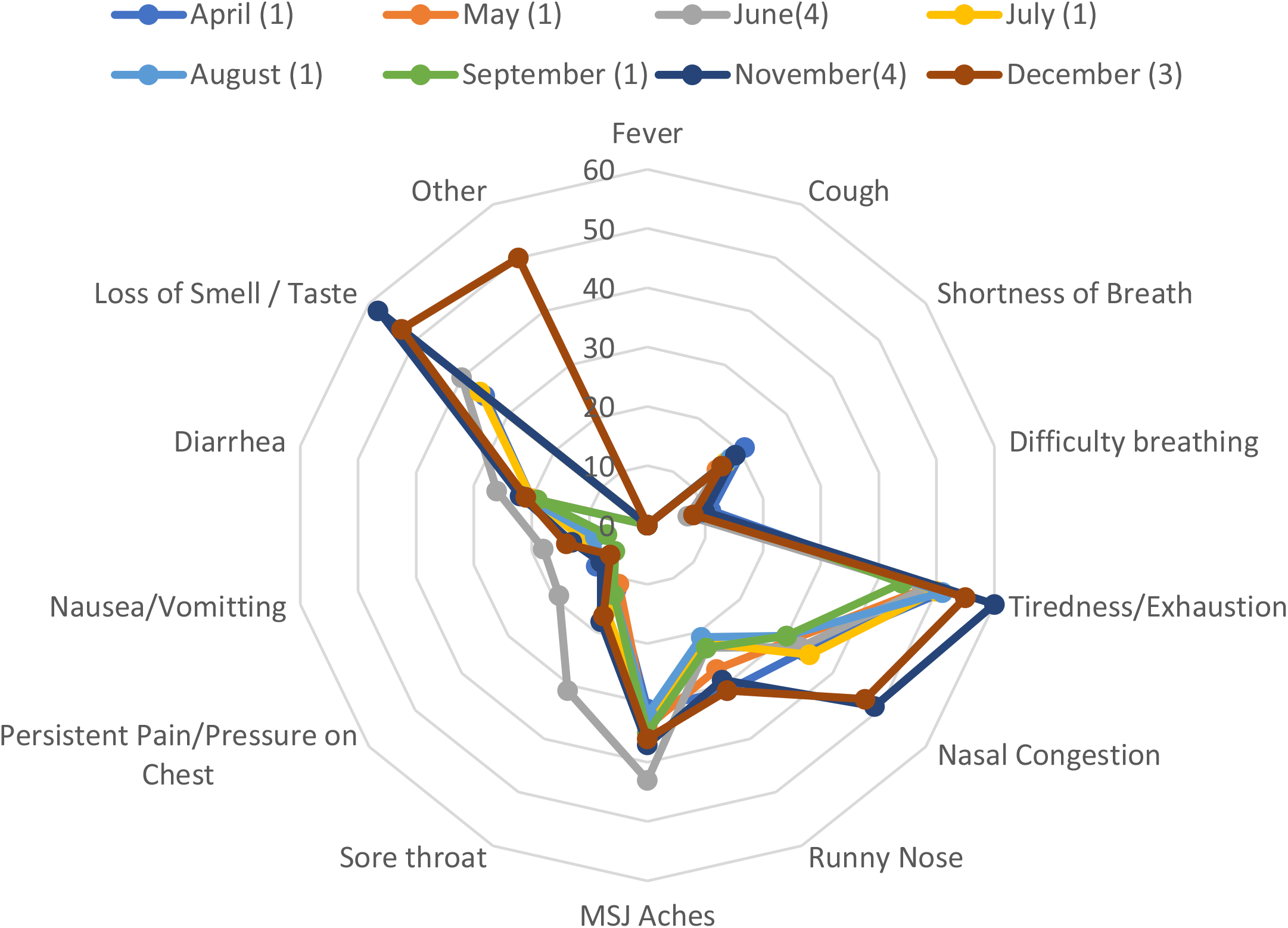
Study Workflow. Phenotype legends: Afebrile (0%), Non-Coughing (0%), Oligosymptomatic (ANCOS) 2. Febrile (100%) Multisymptomatic (FMS) 3. Afebrile (0%) Coughing (100%) Oligosymptomatic (ACOS), 4. Oligosymptomatic with additional self-described symptoms (100%; OSDS) and 5. Olfaction / Gustatory Impairment Predominant (100%; OGIP).

**Table 1.**
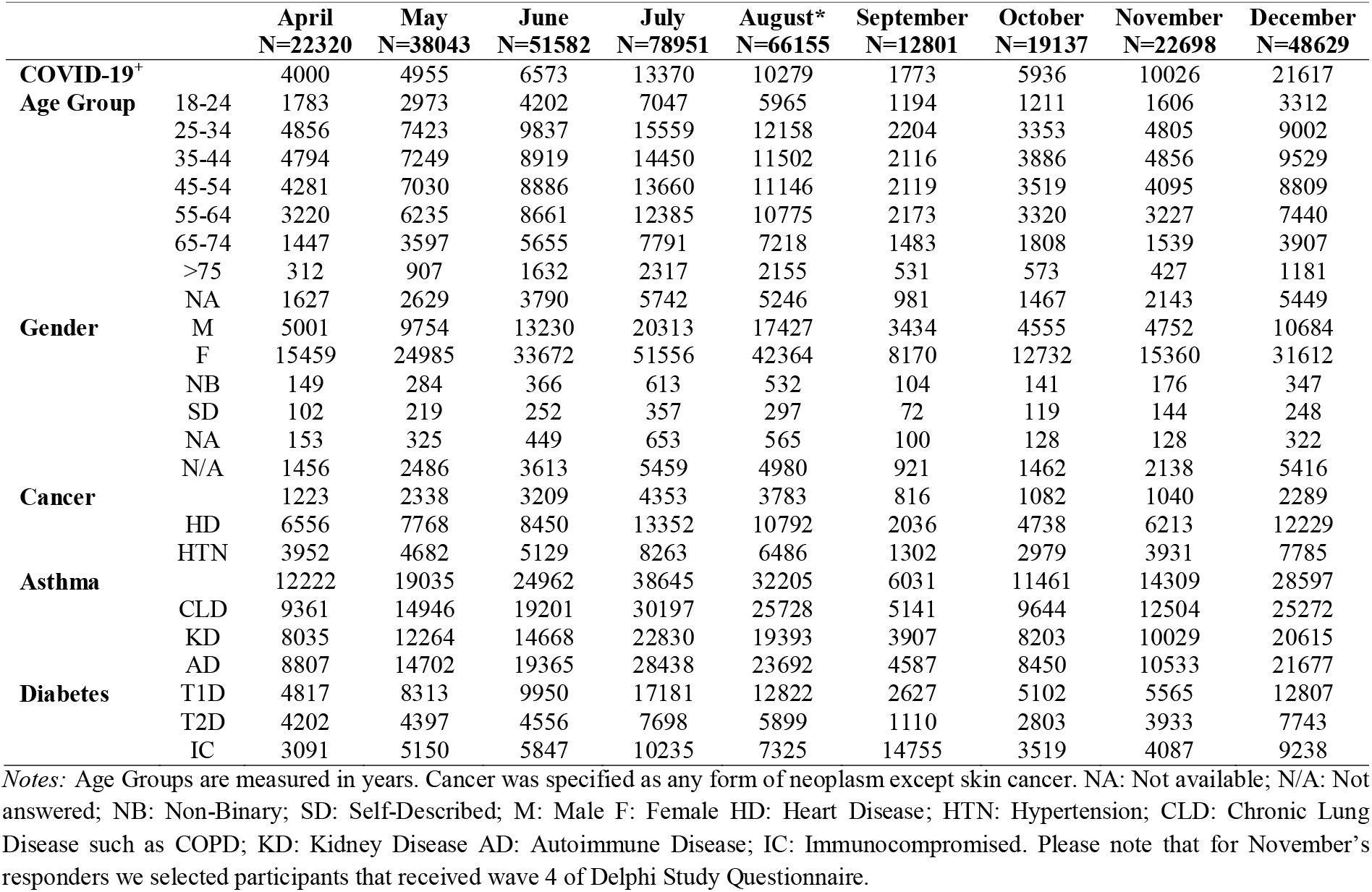
Demographics per month of the study population

**Table 2.** Cluster composition and symptom-based prediction vs. COVID-19^-^ (controls) – August

|  | **N** | **AUC** | **p-value** | **95% CI** |
| --- | --- | --- | --- | --- |
| ANCOS (1) | 2506 | <0.5 | NA | NA |
| FMS (2) | 2266 | 0.963 | <0.001 | 0.961 – 0.965 |
| ACOS (3) | 2060 | 0.737 | <0.001 | 0.729 – 0.746 |
| OSDS (4) | 1013 | 0.777 | <0.001 | 0.762 – 0.792 |
| OGIP (5) | 2434 | 0.983 | <0.001 | 0.982 – 0.984 |

*Notes:* Five COVID-19 syndromes were identified in August: 1. Afebrile (0%), Non-Coughing (0%), Oligosymptomatic (ANCOS) 2. Febrile (100%) Multisymptomatic (FMS) 3. Afebrile (0%) Coughing (100%) Oligosymptomatic (ACOS), 4. Oligosymptomatic with additional self-described symptoms (100%; OSDS) and 5. Olfaction / Gustatory Impairment Predominant (100%; OGIP). AUC: Area Under Curve.

### 4.2. Phenotype extraction

Based on the latent structures between symptom data and disease duration, five COVID-19 syndromes were extracted from August’s 61.165 responders:

1. Afebrile (0%), Non-Coughing (0%), Oligosymptomatic (ANCOS).
2. Febrile (100%) Multisymptomatic (FMS).
3. Afebrile (0%) Coughing (100%) Oligosymptomatic (ACOS).
4. Oligosymptomatic with additional self-described symptoms (100%; OSDS).
5. Olfaction / Gustatory Impairment Predominant (100%; OGIP).

**Figure 2 – 6 and Supplementary Tables 2 – 6** present the symptom composition of each phenotype and its temporal evolution.

**Figure 2 – 6.**
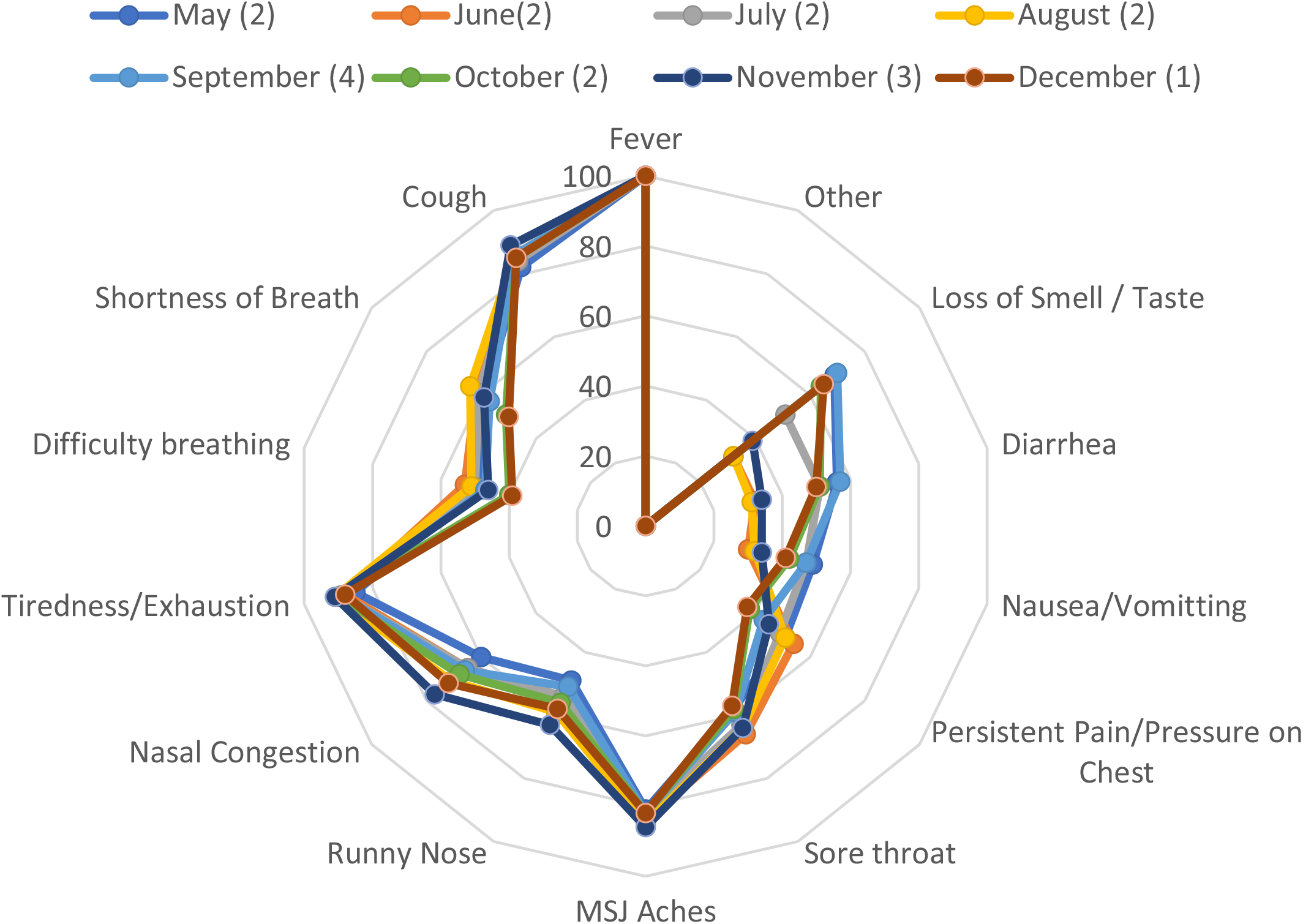
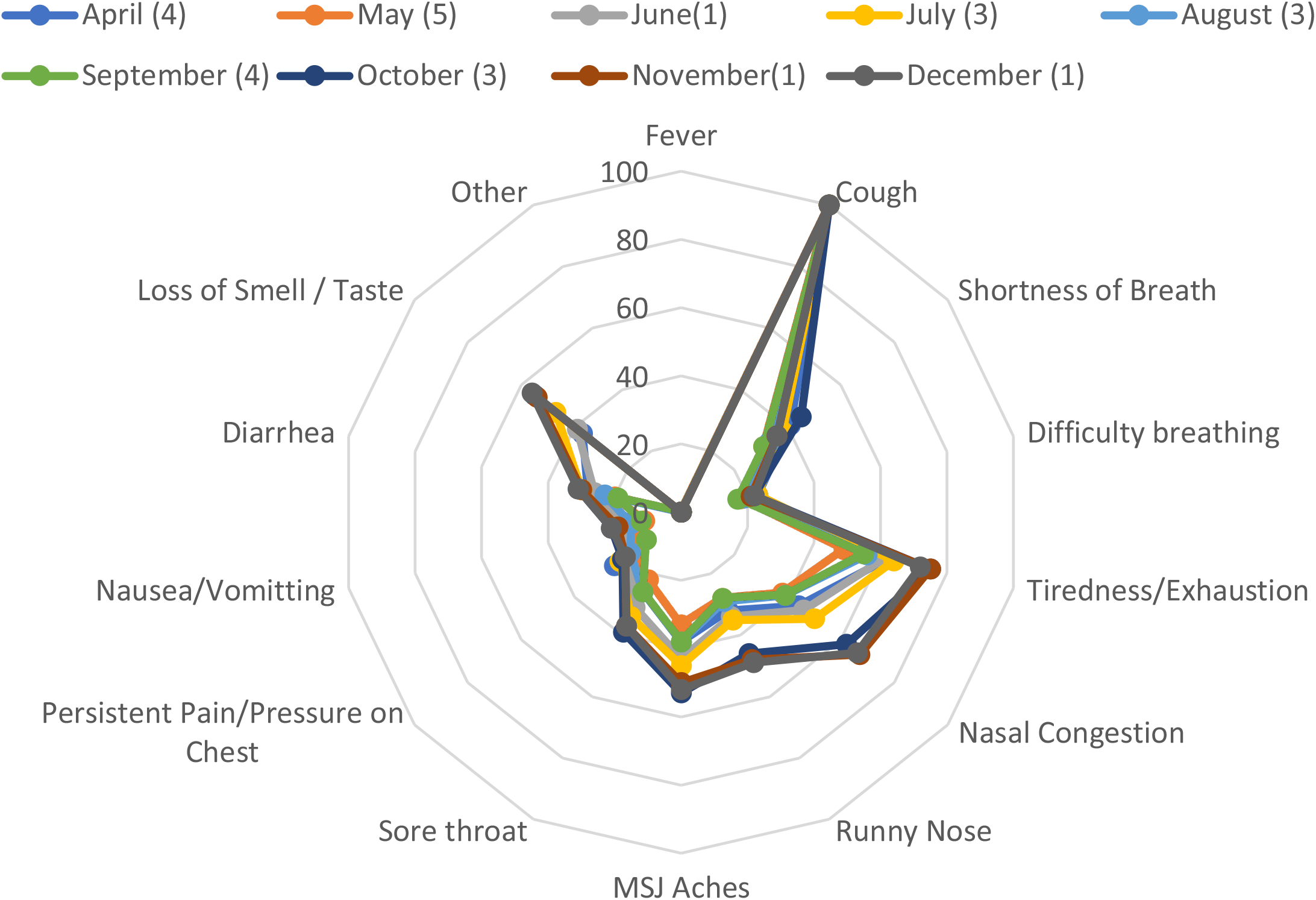
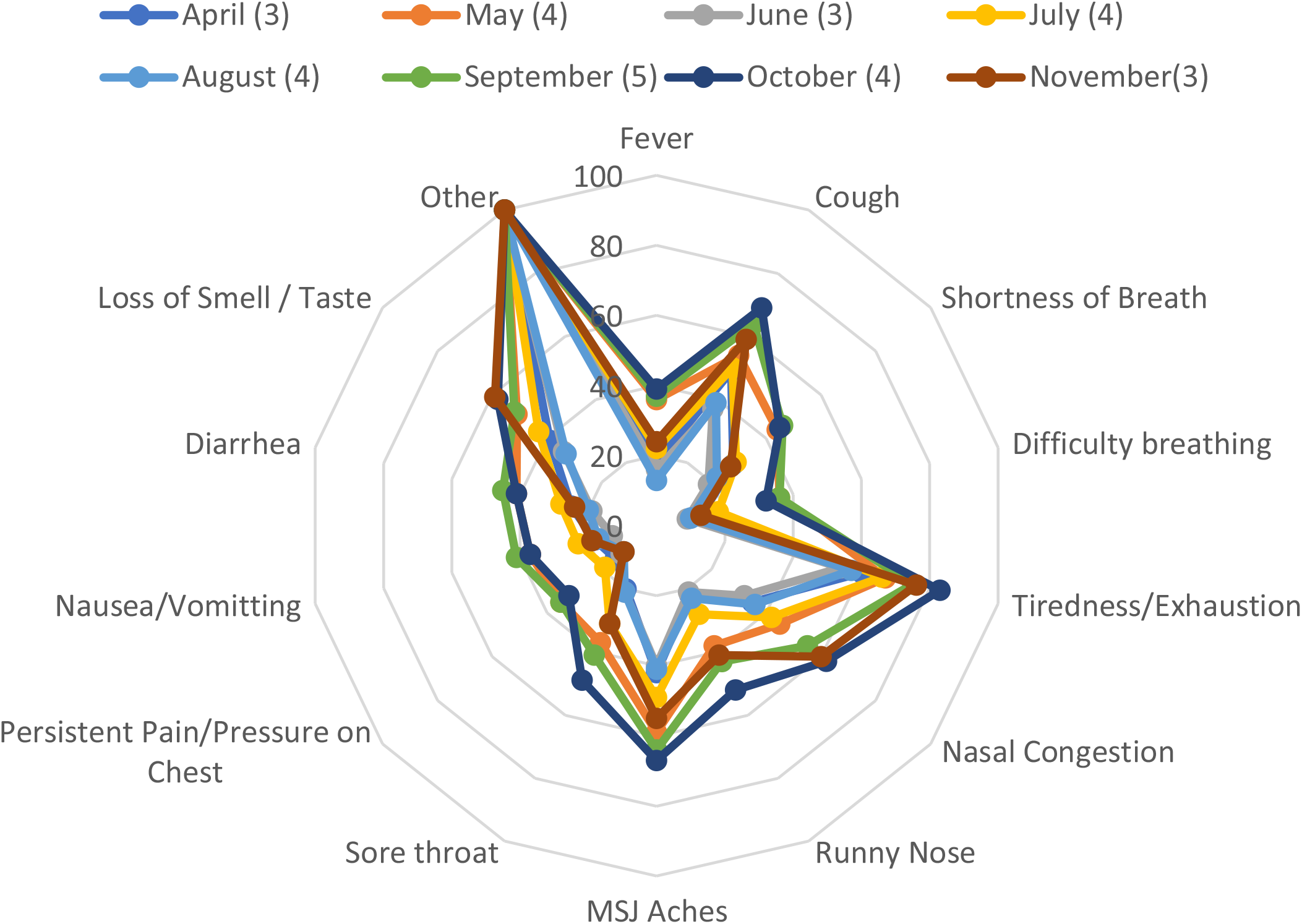
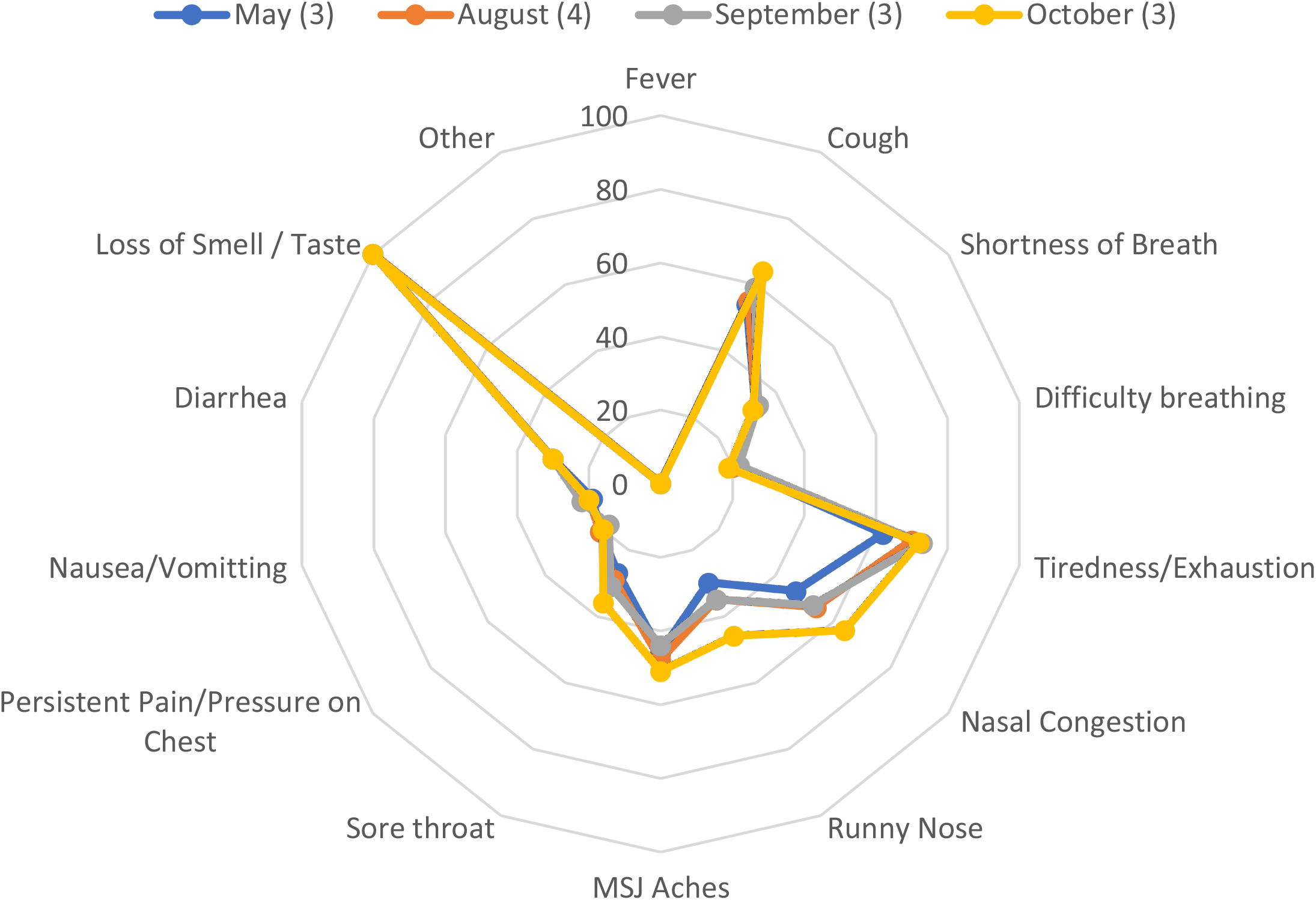
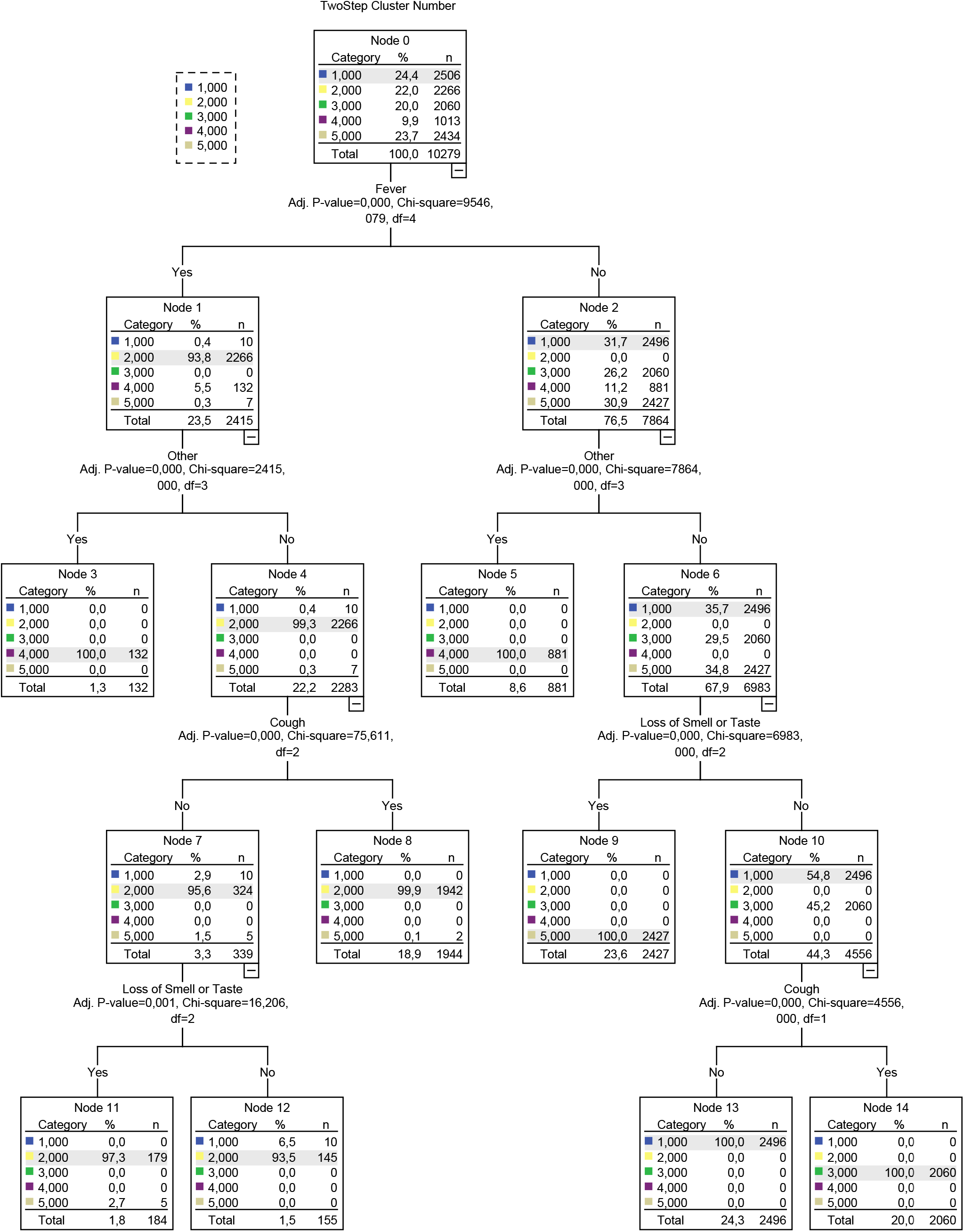
Rose charts and heatmaps presenting the temporal relationships between phenotypes and symptoms. Phenotype legends: Afebrile (0%), Non-Coughing (0%), Oligosymptomatic (ANCOS) 2. Febrile (100%) Multisymptomatic (FMS) 3. Afebrile (0%) Coughing (100%) Oligosymptomatic (ACOS), 4. Oligosymptomatic with additional self-described symptoms (100%; OSDS) and 5. Olfaction / Gustatory Impairment Predominant (100%; OGIP).

### 4.3. Validation and further characterization

Repeating the multiple correspondence and cluster analyses per subsequent and preceding month (April – December), resulted in the validation of each phenotype as follows:

a. ANCOS and OSDS emerged in 10/10 months
b. MFS and ACOS emerged in 9/10 months
c. OGIP emerged in 4/10 months.

Based on the most salient symptoms, decision trees were subsequently constructed (**Figure 7**), providing a structured approach in identifying each phenotype.

**Figure 7.**
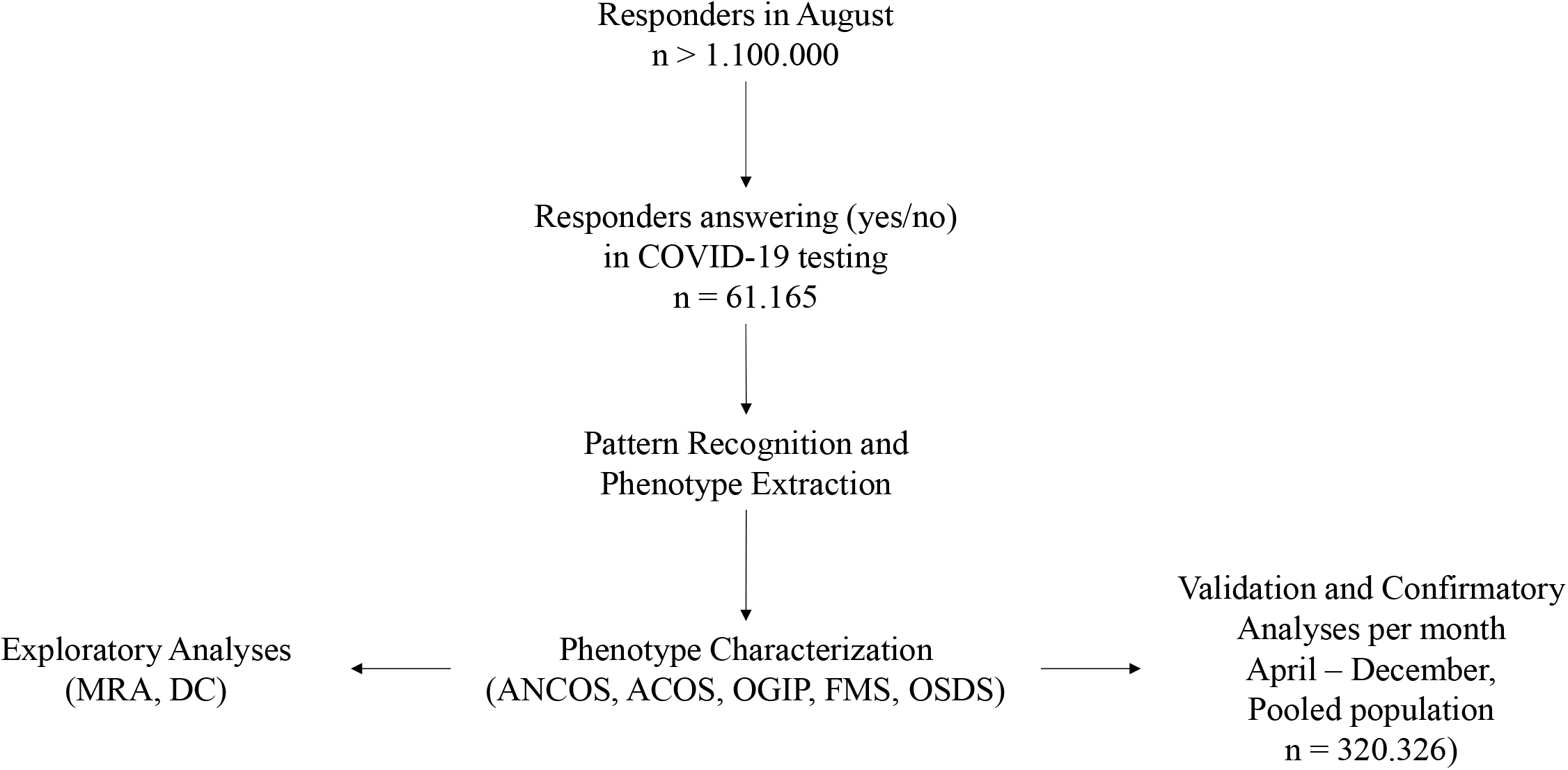
Decision Tree developed using the QUEST algorithm. The decision tree’s branches are based on splits, i.e. variables that were selected based on a chi-square test-determined p-value. The dependent variable for this analysis was a cluster number, represented as a nominal categorical variable with 5 levels corresponding to the 5 clusters.

Further characterization of these 5 phenotypes was achieved via identifying invariant symptoms between April – December (**Supplementary Files / sTables 1a – 1e**); based on these observations and the results of the Shapiro – Wilk tests:

a. ANCOS was characterized by general malaise in the absence of fever and upper respiratory tract symptoms.
b. ACOS was characterized as a mainly afebrile upper respiratory tract viral infection.
c. FMS was a more typical, febrile syndrome covering respiratory and gastrointestinal (GI) manifestations.
d. OGIP, the most invariant syndrome, was characterized by the absence of fever and diarrhea.
e. OSDS did not typically include symptoms of pain or pressure on the chest, nor difficulty in breathing.

Interestingly, the implementation of “Headache” in December as a standalone question resulted in a decomposition of the OSDS phenotype. This is further exemplified by the comparison between text-mined headache as a symptom in August (10% of OSDS) versus a 3-fold increase in prevalence when asked directly in December.

Multiple nominal regression of comorbidities, adjusted for age group and gender, revealed several statistically significant associations (**Supplementary Table 2**). Notably, a history of asthma and chronic lung disease were abortive comorbidities for certain phenotypes (ANCOS, ACOS, OGIP for asthma and additionally OSDS for chronic lung disease). Gender and age group did not display sequentially consistent associations with any phenotype.

## 5. Discussion

In our study, five distinct COVID-19 phenotypes were identified, characterized with respect to comorbidities and demographics, and validated via a data-driven, pattern recognition approach. The presence or absence of fever, cough, olfactory / gustatory dysfunction, and atypical symptoms defined these phenotypes as their primary features. The concept of symptom invariance was subsequently used to further determine their stability regarding symptom composition, indicating that the olfactory / gustatory predominant phenotype OGIP was the most invariant, i.e. the most stable, across the 4 months that it emerged in. Finally, while several comorbidities such as heart disease and diabetes were associated with the risk of manifesting specific phenotypes, other comorbidities such as asthma were found to be abortive.

After its initial identification as a novel pneumonia, the increasing numbers of COVID-19 cases began to outline a spectrum, rather than a linear progression from mild viral infection to a severe one (23). The recognition of COVID-19’s heterogeneity however was initially limited within the setting of treatment response or severity phenotypes (24), (25), while the heterogeneity of non-severe cases or those lacking a salient respiratory aspect was not addressed. Even within the concept of point care phenotyping however, phenotypes similar to those identified in our study have been described by independent studies. Bayesian approaches have identified phenotypes corresponding to FMS, OGIP and ACOS in the clinical setting, and included them in diagnostic algorithms (26).

In our cohort, a similar diagnostic rule emerges and recurs in monthly aggregated data, corresponding to the phenotypes identified here (**Figure 2**).

The importance of phenotyping COVID-19 outside the initially severe or point of care spectrum becomes evident when examining previous iterations of diagnostic criteria: fever and respiratory symptoms were initially the only manifestations considered relevant in defining cases (27). One of the largest studies on initially asymptomatic or non-respiratory symptom (NRS) phenotype of COVID-19 patients has shown that this approach may miss a portion of active cases that may subsequently convert to severe manifestations (28). Our findings support this concept, with NRS overlapping with the OGIP, OSFS and ANCOS phenotypes.

As previous research has suggested, comorbidities were found to be independent predictors of COVID-19 phenotypes, even after adjustments for age group and gender (**Supplementary Table 2, A1 – A5**). As a general rule, two broad categorizations of comorbidities can be inferred: those that can intertwine with the pathophysiology of SARS-CoV-2, such as diabetes (29) and heart disease (30), and those where a treatment effect may restrict phenotype manifestations.

In this light, several noteworthy associations include comorbid asthma and chronic lung disease, which appear to reduce the risk of manifesting the FMS phenotype (**Supplementary Table 2, A2**). This seemingly paradoxical relationship has been previously explored in the literature, and mainly attributed to the protective effects of inhaled corticosteroids (ICS); Specifically, while their use may lead to quiescent type I/III interferon responses, the concomitant downregulation of ACE2 and TMPRSS2 may restrict SARS-CoV-2 from entering pneumonocytes (31,32). As our data are limited regarding medication use and the specific respiratory disease of each responder, we cannot safely attribute the associations observed to ICS usage.

Based on current literature, ICS treatment plausibly presents a potential phenotype abortive effect in otherwise vulnerable populations such as asthma patients (33). Recent evidence on the efficacy of ICS as ad hoc COVID-19 treatments provide further support for this concept (34).

In a similar fashion, the phenotype abortive effect of cancer as a comorbidity could reflect yet another treatment rather than disease effect. Such an effect may account for this association, considering that several anti-cancer treatments are undergoing trials as repurposed COVID-19 treatments (35). As no data were collected on primary tumors, staging and treatment (36) due to the nature of the survey, this hypothesis cannot be scrutinized further.

### 5.1. Limitations and Strengths

The results of our study should be interpreted within the context of their limitations. As a survey administered via Facebook, our source data incur the corresponding selection bias. This however is potentially balanced by the large sample size of the final cohort, and represents the single largest study of its kind. Survivor bias is also inherently present in our study, considering that responders are unlikely to have severe COVID-19 at the time of survey administration. The lack of follow-up data correspondingly precludes that phenotype shifts (e.g. ANCOS or OSDS to FMS) cannot be explored. Another important consideration is that OSDS inevitably absorbs symptoms not originally covered by the initial study iterations and is correspondingly decomposed when these symptoms are identified and added. A prime example of this case is headache as symptom; when left to the discretion of the responder, it might not be evaluated properly as a feature (38). This paradigm becomes evident by the discordance between text-mining (April – November data, 10% of OSDS) vs. asked directly (i.e. 30% in all phenotypes and decomposition of OSDS as a “pure” phenotype). Comorbidities, reported by majority as broad categories, cannot be safely considered in strict interpretations as to their associations with phenotypes. Finally, as gender categories beyond male/female are underrepresented in the monthly samples, they cannot be safely used to extrapolate their contribution on clinical phenotypes. These intrinsic caveats of the study are inherited from the broader structure of the data, and the post-hoc extraction of a data subset for a specific concept (i.e. data driven phenotyping of COVID-19 syndromes).

The main strength of the study was the determination and retro- and anterograde validation of COVID-19 syndromes in the largest community sample to date. The phenotypes we uncover solidify phenotypes previously described by independent studies, and furthermore provide the basis and tools for the development of utilizable diagnostic rules. One of the most important concepts explored here is that the febrile respiratory phenotype represents a lesser portion of COVID-19 phenotypes in the community, a finding that should be considered both in epidemiological profiling and healthcare provision. Our findings support the concept of symptom-based phenotypes of COVID-19, that remain distinct within 9-12 days from first symptom onset. The existence of phenotypes rather than severity strata may further explain the low diagnostic accuracy achieved by rule in or rule out algorithms based solely on symptoms, without accounting for their dependency and intercorrelations, even between different systems (i.e. GI and respiratory). In order to utilize our findings in the clinical setting and make them available to other researchers, we have developed an online application that calculates the symptom-based logistic probability *P*_*x*_ (Available from: http://se8ec.csb.app). In this community-based sample, febrile respiratory disease was infrequent compared to atypical presentations within a range of 9 – 12 days from symptom onset; this finding may be critical in current epidemiological surveillance and the development of transmission dynamics concepts.

## Supporting information

Heatmap 1

Heatmap 2

Heatmap 3

Heatmap 5

Heatmap 6

sTable 1

## Data Availability

The analyses and all related files are available upon request.

https://cmu-delphi.github.io/delphi-epidata/symptom-survey/

## Acknowledgements

This study was made possible by the collaboration with the Carnegie Melon’s Delphi Group Survey Data and Facebook’s Data for Good initiative. The authors would like to thank Alex Reinhart and Wichada Le Motte-Kier for their impeccable work in organizing the survey and coordinating the investigator assemblies; Kathryn Rivard-Mazaitis for her invaluable help with the survey data access; Kelsey Mulcahey and Stewart Tansley from Facebook, for facilitating the collaboration with Delphi Group, and their work on the Facebook study; Athanasia Kefala for her help with the rose charts.

